# High confidence and demand for hepatitis E vaccine during an outbreak in Bentiu, South Sudan: A qualitative study

**DOI:** 10.1101/2024.06.25.24309497

**Authors:** Aybüke Koyuncu, Kinya Vincent Asilaza, John Rumunu, Joseph Wamala, Priscillah Gitahi, Zelie Antier, Jetske Duncker, Patrick Nkemenang, Primitive Gakima, Melat Haile, Etienne Gignoux, Manuel Albela, Frederick Beden Loro, Duol Biem, Monica Rull, Andrew S Azman, Iza Ciglenecki, Robin Nesbitt

## Abstract

**Introduction:** In 2021 in response to an outbreak of hepatitis E in Bentiu internally displaced persons camp the South Sudanese Ministry of Health with support from Médecins Sans Frontières implemented the first-ever mass reactive vaccination campaign with HEV239 (Hecolin; Innovax, Xiamen, China). As part of an evaluation of the feasibility of hepatitis vaccination as part of an epidemic response, we conducted qualitative research to assess knowledge, attitudes, and practices related to hepatitis E and the hepatitis E vaccine.

**Methods:** We conducted 8 focus group discussions (FGDs) with community leaders, the general population of vaccine-eligible adults, vaccine-eligible pregnant women (vaccinated and non-vaccinated), and healthcare workers. FGDs were separate by gender and were audio recorded, transcribed, and translated to English by trained research assistants. Two coders used inductive thematic analysis to organize emergent themes.

**Results:** Data were collected in November 2022. Most individuals had personal experiences with hepatitis E. Hepatitis E was perceived as a dangerous disease, and almost everyone was knowledgeable about transmission pathways. Participants believed children, pregnant women, and the elderly were the highest risk groups. Participants frequently made requests for additional hepatitis E vaccination campaigns and expanded eligibility criteria for vaccination. The primary barriers to vaccination were practical issues related to being away from the camp at the time of the campaign, but participants shared that some in the community were unvaccinated due to fears about injections, social pressure, misinformation about side effects such as infertility, concerns about why some groups were eligible for vaccination and not others (e.g. young children), and a lack of information about the vaccine/vaccination campaigns.

**Conclusion:** Personal experiences with hepatitis E illness, perceived severity of illness, and confidence in organizations recommending the vaccine were drivers of high demand for hepatitis E vaccines in the first-ever use of the vaccine in an outbreak setting.

## Introduction

Hepatitis E genotypes 1 and 2 cause over 3 million symptomatic cases of acute viral hepatitis each year with case fatality risk as high as 65% among pregnant women [1–3]. Large outbreaks of hepatitis E virus (HEV) genotypes 1 and 2 occur due to fecal contamination of drinking water, and at least 1 large-scale outbreak with over 5,000 suspected hepatitis E cases has occurred in every decade since 1988 [4]. While outbreaks occur in resource-limited settings throughout Africa and Asia, refugees and internally displaced persons are at heightened risk for outbreaks due to overcrowding and poor access to safe water, sanitation, and hygiene (WASH) [1,5].

One potential tool for HEV outbreak control is a recombinant vaccine, HEV239 (Hecolin; Innovax, Xiamen, China), which has been proven to be safe and efficacious in individuals 16-64 years old [6]. The vaccine is licensed only in China and Pakistan and has not been recommended by the World Health Organization (WHO) for routine use in HEV-endemic countries, largely due to a lack of epidemiologic data in the general population [1]. Despite a lack of data to justify routine use of the vaccine, in 2015 WHO recommended the vaccine be considered as a strategy to mitigate or prevent outbreaks [1]. While the current vaccine’s 3-dose schedule given across 6 months is not ideal for rapid deployment in outbreak settings [7], multiyear protracted HEV epidemics suggest the potential for even a 6-month regimen to substantially prevent morbidity and mortality, particularly among high-risk groups such as refugees and pregnant women.

In Bentiu IDP camp, hepatitis E cases have been reported since 2014 with large outbreaks occurring in 2015-2016 and again in 2019. Between October 2014 and April 2022 there were 2,227 confirmed cases of hepatitis E in Bentiu camp. In response, the South Sudanese Ministry of Health (MOH) in partnership with MSF implemented the first-ever vaccination campaign for hepatitis E in the context of an outbreak beginning in March 2022. The campaign targeted individuals 16-40 years old residing in Bentiu camp, including pregnant women. The campaign took place in three rounds in March, April and October 2022 [8]. In parallel, MOH and MSF implemented operational research to understand the effectiveness, safety, and feasibility of vaccination against hepatitis E. As part of this research, a vaccination coverage survey was conducted immediately after the third and final vaccination round. Coverage with at least one dose of the vaccine in the target population was 86% (95% CI: 84-88), and the most frequent reasons for non-vaccination were physical absence (60%) and fears and concerns (18%) [9]. Here we report findings from the qualitative component of the study aimed at understanding community perspectives on hepatitis E, hepatitis E vaccine, and barriers and facilitators of vaccine uptake.

## Materials and methods

### Setting

Bentiu internally displaced persons (IDP) camp was established in 2014 as a Protection of Civilians site at a United Nations Mission in South Sudan base in response to conflict in Unity State. The camp has expanded since 2014 and the total camp population in 2022 was approximately 112,000 residents.

### Participants

We recruited community leaders, the general population of vaccine-eligible adults(i.e. 16-40 years, resident of Bentiu IDP camp), vaccine-eligible pregnant women (vaccinated and non-vaccinated), and healthcare workers to participate in focus group discussions (FGDs). The general population was recruited using convenience sampling in each camp sector from individuals already participating in research on vaccine coverage while pregnant women were recruited from individuals already participating in research on vaccine safety. Community leaders and medical staff were approached separately for recruitment. We asked the Camp High Commission to recommend community leaders and asked managers at health facilities to recommend medical staff working with hepatitis E patients.

### Data collection

Data were collected in November 2022. We selected the Behavioral and Social Drivers of Vaccination (BeSD) framework [10] *a priori* to develop FGD guides and organize emerging themes for knowledge, attitudes, and practices about hepatitis E, perceptions about the hepatitis E vaccine, and barriers and facilitators of hepatitis E vaccine uptake. The BeSD framework is a theory-based framework for understanding individual and community-level drivers of vaccination uptake that is organized into four domains: thinking and feeling, social processes, motivation, and practical issues (Fig 1).

**Figure 1.**
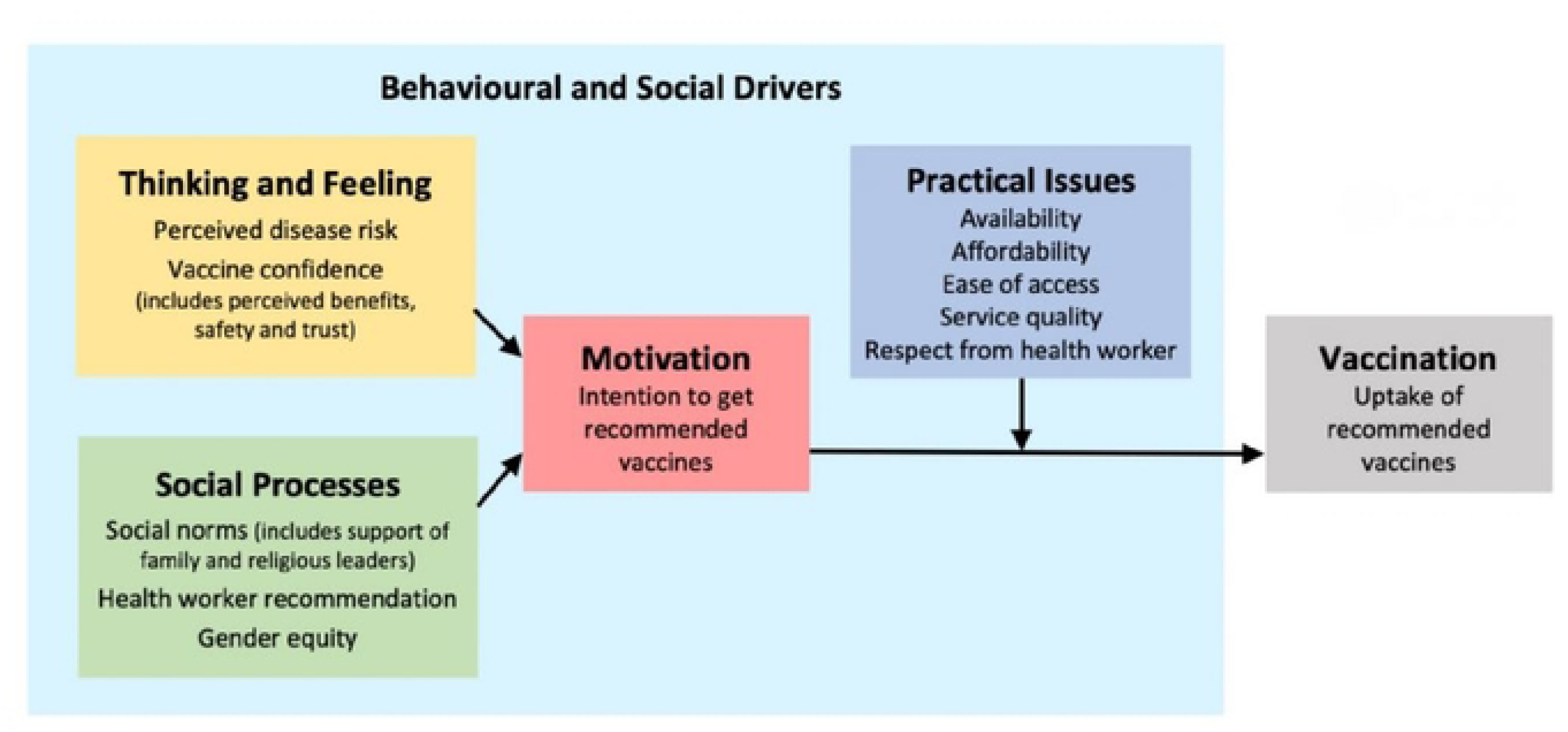
The behallloral andsocial drivers of vaccination (BeSO) framework. Based on the BeSO working group (10) and adapted from Brewer et al. [11].

FGDs were conducted separately by population type and gender except for healthcare workers. FGDs were conducted in Nuer or English and were audio recorded, transcribed, and translated to English.

### Analysis

Within domains of the BeSD framework, two coders (AK and RN) used inductive thematic analysis to iteratively read transcripts and code the text in the data. Emergent key themes and codes were entered into a codebook, and codebooks and the categorization of data were compared for 2 FGDs. We used a negotiated agreement approach to assess intercoder reliability [12], with differences in coding adjudicated by discussion and consensus between coders. Once the emergent codes and themes were standardized, both coders independently coded the text in the remaining data using Nvivo software (QSR International, Melbourne, Australia). Once the emergent codes and themes were standardized, both coders independently coded the text in the remaining data using Nvivo software (QSR International, Melbourne, Australia).

### Ethics

Ethical approval was granted from the MSF Ethical Review Board (ERB) and by the South Sudan Ministry of Health Research Ethics Board as part of the study protocol titled: “Effectiveness, safety and feasibility of recombinant hepatitis E vaccine HEV 239 (Hecolin) during an outbreak of hepatitis E in Bentiu, South Sudan” (MSF ERB #2167 and RERB-MOH # 54/27/09/2022). All participants provided verbal informed consent before participating in a FGD.

## Results

We conducted 8 FGDs with 6-8 participants each (48-64 total participants): 2 FGDs with community leaders, 2 FGDs with the vaccinated general population, 2 FGDs with the unvaccinated general population, 1 FGD with healthcare workers, and 1 FGD with pregnant women. Participants from the general population were aged 18-40 while community leaders and healthcare workers included participants outside of the vaccine-eligible range. Emergent themes were consistent by participant type and gender.

### Thinking and feeling

All population types were very knowledgeable about hepatitis E symptoms, transmission pathways, and prevention methods. The most frequently mentioned symptoms associated with hepatitis E were yellow eyes, fever, and dark urine. Some participants also mentioned yellow or “light” skin and general weakness of the body. Cleanliness and WASH were frequently reported as key prevention strategies. Many respondents felt it was difficult to prevent HEV infection with vaccine alone because of the crowded conditions in the camp:

> *“All toilets are full and we are trying to advice people to be vaccinated yet the problem of toilets is very dangerous one.”*
>
> — *Male community leader*

Most participants had high perceived risk of being infected with hepatitis E and perceived hepatitis E as a dangerous and serious infection. Hepatitis E was perceived as more severe in comparison to other infectious diseases such as malaria:

> *“HEP E is dangerous compared with malaria; malaria can be treated but HEP E can take long to be treated.”*
>
> — *Male community leader*

Other reasons for perceiving hepatitis E as more serious compared to other infections included potential impacts of infection on family finances (e.g if the breadwinner is infected), the potential for infection to be fatal, and knowledge that there is no existing treatment. Many individuals described traditional healing methods for hepatitis E that were used in their villages and also in the camp which involve beating the infected person with a hot metallic stick.

> *“During the village life, when you are HEP E positive, the only thing people do is beat you with metallic stick put on fire for you to get well, but now we have the vaccine given by MSF freely without paying any single money. when you are beaten with that thing put on fire, you will feel pain at the same time.”*
>
> — *Pregnant female community member*

Most individuals had personal experiences with hepatitis E including being infected themselves or having a family member, neighbor, or community member that had been symptomatically infected:

> *“In the past, during the time of village life, my sister was infected with HEP E, because there were no vaccine, she died, but now the vaccine is brought by MSF…”*
>
> — *Pregnant female community member*

> *“Community leaders like the vaccine because they have seen the how worse is HEP E in the past and it infected some of them, some of them were involved during the campaign too.”*
>
> — *Female community leader*

Children, pregnant women, and the elderly were consistently identified as high-risk groups for HEV infection and/or severe outcomes. Children were perceived as being at high-risk because they play in dirty areas in the camp, are more likely to practice open defecation and don’t know how to take care of themselves, have weak immune systems, and because they forget instructions from parents (e.g. to not play in dirty areas) and can’t communicate symptoms when they are ill. Many participants questioned the eligibility criteria for vaccination and requested that future campaigns include children and elderly in the target population:

> *“In this camp, many people are infected, but mostly children. My question is that, why MSF is not bring the vaccine for children? And this disease is affecting children serious through dirty water and dirty playing in dirty environment.”*
>
> — *Male community leader*

Participants were aware of the risk of severe outcomes among pregnant women, and the elderly were perceived as a high-risk group for severe outcomes due to weakened immune systems.

> *“When this disease infect the pregnant mother, its not easy for such a mother to survived.”*
>
> — *Healthcare worker*

Participants had high confidence in vaccine effectiveness and benefits, vaccine safety, and the organizations recommending the vaccine. Most participants believed it was safe for women to receive the vaccine during pregnancy, though some unvaccinated males acknowledged not knowing if the vaccine was safe for pregnant women. Many participants reported positive impacts of the vaccine campaign such as a perceived reduction in the incidence of hepatitis E. Participants mentioned some community members refused the vaccine due to low knowledge about the importance of the vaccine and rumors about vaccine safety circulating in the community (e.g. fears about infertility following vaccination), but did not report being personally concerned about benefits, safety, or side effects. Some participants reported mild side effects after vaccination such as headaches or fever that cleared in a few days. Some vaccinated individuals reported fear of injections as a personal barrier to vaccination that they overcame. Others mentioned concerns about eligibility as a reason for non-vaccination among community members:

> *“Some people refused the vaccine because they said that why MSF is vaccinating some group and leave others to be killed by the disease? Some refused for that reason.”*
>
> — *Male community leader*

### Social processes

Most individuals were in support of vaccination and had been vaccinated. Individuals in the general population perceived that most of their community and religious leaders were in support of vaccination, and had high respect for healthcare workers recommending and providing vaccines:

> *“We trust also the people who offered to us the vaccine because they are the members of this community”*
>
> — *Vaccinated female community member*

Trusted relationships with vaccinators and organizations promoting the vaccine were influential in the decision to get vaccinated. Participants reported trusting the vaccine and vaccinators because the vaccine was brought by doctors, the vaccine was brought by MSF, and because the government permitted the vaccination campaign:

> *“As other people talk about it before, this vaccine is very saved for the community because it was brought by doctors from MSF, if we get it from other sources, we will never trust the vaccine at all, if bush men come and tell us about this vaccine, we will never accepted to be vaccinated.”*
>
> — *Female community leader*

Though decision-making was not mentioned explicitly in all FGDs, some participants mentioned the final person to tell whether family members should be vaccinated or not was either the father or mother of the family. Some participants mentioned pressure from other community members to refuse vaccination and circulating rumors about side effects such as infertility and an increased risk of getting other diseases.

### Motivation

Individuals were highly motivated to get vaccinated. There were frequent requests for additional hepatitis E vaccination campaigns in the future and expanded age and geographic eligibility in future campaigns:

> *“My question is about those who are living outside the camp, is there any mean for this to get the vaccine? Remember these people are part of this community.”*
>
> — *Male community leader*

### Practical issues

Key facilitators of vaccination were that the vaccine was available free of charge and that vaccinators went to all areas of the camp. The primary barrier to vaccination among unvaccinated adults was being away from the camp at the time of the campaign:

> *“I was not vaccinated because I was far away from the camp during the vaccination, so I was unlucky, my appeal to MSF is if there is still vaccine remain, we should be considered.”*
>
> — *Unvaccinated female community member*

Some participants also reported a lack of information about the vaccine and the second and third rounds of the vaccination campaign as a barrier to vaccine uptake.

## Discussion

Our study identified high demand for hepatitis E vaccines among residents of Bentiu IDP camp, following the first-ever use of the vaccine in an outbreak setting. Personal experiences with hepatitis E illness, the perceived severity of illness, and high confidence in the healthcare providers and organizations recommending and providing the vaccine were key facilitators of vaccine uptake. Perceptions about hepatitis E and the hepatitis E vaccine in our study were consistent by vaccination status, gender, and population type, with the consistency of key themes suggesting data saturation. For example, even unvaccinated adults in our study reported predominantly positive perceptions about the vaccine and asked for additional opportunities to be vaccinated. While improvements to comprehensive WASH infrastructure are needed for long-term control of hepatitis E and other pathogens as noted by community members, we identified high demand for hepatitis E vaccines as a tool for prevention and control of hepatitis E outbreaks.

During the second and third rounds of the campaign, the vaccine was offered to anyone within the target group (16-40 years old and residence in Bentiu IDP camp), regardless of whether they received previous doses or not (8). The campaign also had a longer duration than most reactive vaccination campaigns: each round of the campaign lasted more than a week, and the final round lasted more than 2 weeks. Despite these attempts to improve vaccine access, the primary self-reported barrier to vaccine uptake was being away from the camp at the time of the campaign. Our qualitative findings are consistent with the quantitative coverage survey conducted in Bentiu and with findings about primary reasons for non-vaccination in preventative and reactive vaccination campaigns with oral cholera vaccines in various settings [13–15]. The population in Bentiu IDP camp, like many other camp settings, is highly mobile and spends long periods of time outside of the camp. Expanding access to vaccines beyond short-duration campaigns can improve vaccine uptake and increase the likelihood of sustainably maintaining coverage in highly mobile populations such as IDPs. Additional research is needed to explore the potential costs and benefits of vaccination strategies such as extending the duration of the campaign, offering the vaccine to all new camp entries, or routinely offering the vaccine in health facilities. The possibility of implementing these strategies for hepatitis E vaccine is further complicated by Hecolin’s bulky, single-dose, pre-filled glass syringes which introduce additional challenges for transport, storage, and waste management.

Children and the elderly were consistently identified as high-risk groups for HEV infection and severe outcomes by the community in Bentiu but were ineligible for vaccination in the campaign. For some individuals, the exclusion of children and the elderly from the campaign contributed to vaccine refusal for themselves. The age target for the campaign (16-40 years) was chosen because the vaccine is not currently registered for use in children younger than 16 years and due to limited doses of the vaccine the upper limit was chosen based on the lower attack rate for hepatitis E among older adults in the camp (8). Notably, perceptions about children as a high-risk group for infection are mirrored in surveillance data on hepatitis E in the camp (68% of confirmed hepatitis E cases based on rapid diagnostic tests between October 2014 and April 2022 were aged 10 years or younger (unpublished data)). Additional research is needed to understand the role of children in HEV outbreaks, the safety and efficacy of the vaccine in children, and the potential value of expanding age-eligibility for the vaccine.

Even amongst unvaccinated participants, we identified few participants with negative views about the vaccine. This may be because the primary barrier to vaccination was being away from the camp. Though not mentioned explicitly by participants in our study, it is also possible that individuals who previously refused the vaccine may have changed their views after witnessing the safety and benefits of the vaccine in their community. Individuals who agreed to participate in this study also may have been more likely to have positive views about the vaccine compared to the general population in Bentiu, and may have been more likely to report positive views during FGDs due to social desirability bias. In an effort to reduce social desirability bias we asked participants to share community perceptions in addition to personal perceptions. Our findings are also consistent with anecdotal data from staff involved in the vaccination campaign as well as results from the coverage assessment which demonstrated high uptake of the vaccine (9). Studies on the acceptability of hepatitis E vaccines are limited, and vaccine confidence can depend on disease-specific knowledge, attitudes, and practices. However, our findings that perceived severity of hepatitis E was a key facilitator for high vaccine confidence and uptake are consistent with qualitative findings from Juba, South Sudan examining drivers of high oral cholera vaccine uptake during a humanitarian crisis [16]. Our findings may not be generalizable to outbreak settings with limited historical outbreaks of hepatitis E or to settings where trusted relationships with MSF and/or the government are lacking. However, our findings underscore the importance of building trust with communities before, during, and after outbreaks as a strategy for building vaccine confidence.

## Conclusion

Hepatitis E is a known and feared disease among the population, and vaccination against it was a widely accepted and sought-after method of preventing it. Cultivating trusted relationships with communities over time can help individuals overcome common barriers to vaccine confidence. Addressing practical issues related to being away from the camp at the time of the campaign can improve coverage in future campaigns.

## Data Availability

Excerpts of the transcripts relevant to the study are available within the paper.

## Acknowledgments

We would like to thank the MSF hepatitis E vaccination and hospital study teams in Bentiu for their support in implementing the campaign and operational research, as well as the study participants for sharing their perspectives and experiences.

